# Progress towards the Ending Preventable Maternal Mortality (EPMM) target for Bangladesh: The role of female education

**DOI:** 10.1101/2020.06.01.20119479

**Authors:** Karar Zunaid Ahsan, Peter Kim Streatfield, Kanta Jamil, Shams El Arifeen

## Abstract

Educational attainment among women is a well-recognized predictor for maternal mortality. Data from nationally representative surveys and the United Nations are used in the analysis for estimating maternal mortality due to improved education status up to 2030. Analysis of data from 2001 and 2010 Bangladesh Maternal Mortality Survey shows that MMR varies considerably by education level. The study shows that during 2011–2030, 15% maternal deaths will be averted due to fertility change (i.e. fewer births) and 24% of the maternal deaths can be averted only by improving the female education levels. However, in order to achieve the Ending Preventable Maternal Mortality (EPMM) target of 59 maternal deaths per 100,000 live births by 2030 for Bangladesh, a further 64% reduction will be required. Factors outside the health sector, like female education, will continue to have an impact maternal mortality in Bangladesh. However, reaching the EPMM target for Bangladesh by 2030 will also require significant investments in maternal health programs, in particular those to increase access to and quality of services.

## Introduction

Maternal ill health and pregnancy-related complications are major contributors to the disease burden affecting women, each year causing approximately one third of a million deaths worldwide, 99% of which are taking place in developing countries [1]. Earlier studies estimated that compared to the number of maternal deaths, burden of injuries during delivery that often leads to permanent disability among women of reproductive age is 30 times higher [2].

A collaboration among individuals and institutions established in 2005, the Countdown to 2015, to stimulate country action by tracking coverage for interventions needed to attain Millennium Development Goals (MDGs) 4 and 5. Bangladesh is one of the only nine out of 75 Countdown countries that are on track to achieve the primary target of MDG 5 by 2015 [3]—the Maternal Mortality Ratio (MMR) in Bangladesh fell from 322 deaths per 100,000 live births (95% CI: 253–391) in 1998–2001 to 194 deaths per 100,000 live births (95% CI: 149–238) in 2007–2010 [4, 5]. This impressive decline in MMR in Bangladesh seems to have been the result of factors both within and outside the health sector—apart from decrease in mortality risk mainly due to improved access to and use of health facilities, a number of favorable socio-demographic changes, including fertility reduction and improved education levels of women of reproductive age, significantly contributed to such decline in MMR [6].

Among the key contributing factors of maternal mortality reduction in Bangladesh reported by Arifeen and colleagues [6], this analysis focuses on the role of female education and attempts to project MMR for 2030 to estimate the impact of rising education levels among women of reproductive age on maternal mortality. The projected MMR levels are compared with the post-2015 development agenda for ending preventable maternal mortality (EPMM) target for Bangladesh [7] and several policy considerations are proposed in this paper.

### Impact of female education on maternal health and mortality

There has been a renewed interest on the effect of social determinants of health, particularly of women’s educational attainment, on maternal mortality [8]. Studies in different countries have established that years of formal education among women are an important predictor for mortality in their own right [9, 10, 11] and affect health service utilization despite adverse family or socioeconomic situations [12, 13].

What are the pathways that education operates through to reduce maternal mortality? The most obvious pathway for education to influence maternal mortality is by making women better informed, primarily through health education programs (e.g. recognition of obstetric danger signs, health practices that may influence safer childbirth procedures), and enabling them to make better choices regarding antenatal, delivery, and postnatal care [14, 15]. In terms of socio-psychological pathway, Caldwell and his colleagues [16] have established how education can have an empowering effect on women by broadening their horizons and making them aware of available opportunities. The final possible distal pathway of influence refers to enhanced opportunity for educated females to seek paid employment, and thus delaying marriage and child birth [17,18] as well as improving access to resources [15], which is crucial to avail services during emergencies and/or from the private sector. Also, a dominant behavioral pathway linking education to fertility is the use of modern, effective contraceptive methods [19] that reduces exposure to pregnancy-related risks when pregnancies are unwanted or known to be of high risk [20].

In Bangladesh, steady improvement in female education has been a key factor in bringing down the MMR in Bangladesh during 2001–2010 [6]. Knowledge of maternal complications varies substantially by education level—only 20% of women with no education had knowledge of four or more life threatening pregnancy complications, compared with 39% among women with secondary complete or higher education [21]. Pregnant women with higher education status remained significantly more likely to choose safe maternal practices (viz. skilled attendance during delivery or delivery in a health facility)—women with secondary or higher education are 13 times more likely to use medically trained personnel for delivery, and have nearly five times lower MMR than women with no education [4, 5]. A 30-year cohort study also demonstrated that educational differentials for maternal mortality were substantial—the odds ratio for more than 8 years of schooling compared with no schooling was 0·30 (95% CI: 0·21– 0·44) for maternal mortality and 0·09 (95% CI: 0·02–0·37) for abortion mortality [22]. This implies that women with secondary incomplete or higher education are 70% less likely to die during childbirth compared with women with no education, and 91% less likely to die due to abortion complications.

Educational attainment is also closely associated with other factors that have direct effect on maternal health and mortality risks—successive rounds of Bangladesh Demographic and Health Survey (BDHS) data showed that during 1993–2011, ever-married women with no education have been older and poorer than the women with any educational attainment [23, 24].

### Secondary school stipend and female education trends in Bangladesh

Since Independence, the emphasis of the Government of Bangladesh’s human resources development priorities was normally placed on basic education, consisting of primary (grades 1–5), junior secondary (grades 6–8), secondary (grades 9–10), higher secondary (also referred to as college education, grades 11–12), and non-formal education. While primary school education in rural areas of Bangladesh is dominated by public and non-government organization (NGO)-run schools, secondary schools are largely non-government or private [25]. Bangladesh has some of the longest-running education transfer programs in the world, beginning with the Food for Education (FFE) program in 1993 that distributed wheat and rice to poor households in disadvantaged rural areas for sending children to school. In 2002, the Primary Education Stipend (PES) program replaced FFE and more than 10 million children received a stipend up to 2013—the medium-term impact assessment of PES indicated improved school enrolments, household expenditures, and calorie and protein consumption in rural Bangladesh [26].

The gender inequality in schooling, particularly at the post-primary level, was pervasive in the early 1990s. Less than one-third of the total enrolees in secondary schools were girls, and only 14% of girls aged 11–16 were enrolled in secondary schools in 1990 [27]. The dropout rate in secondary schools was also more than 60% with girls than boys in 1990 [27]. In order to address the gender gap in secondary education, monetary incentives to girls in secondary school to cover education expenses in the form of a stipend program was piloted in 1982 by Bangladesh Association for Community Education (BACE), a national NGO with USAID financial assistance under the supervision of the Asia Foundation [28, 29]. The experience of this pilot was found to be highly successful—girls’ secondary enrolments increased from an average of 8% to 14% in some project areas and dropout rates fell from 15% to under 4% [30]. In the light of the pilot’s success, the Female Secondary School Stipend Program (FSSP) was expanded nationally in 1994 with government and donor support [29]. The amount of the monthly stipend varied between US$ 12 for grade 6 and US$ 36.25 for grade 10, and the conditions for receiving the stipend were to attend recognized institutions, remain unmarried, maintain at least 75% attendance, and secure at least 45% marks in the annual examinations [25, 28]. Reviews of FSSP indicated that aggregate female student secondary enrolment increased by 111% during 1994–1999 [28]; years of education for eligible girls increased by an average of 14%–25% [25]; and delayed age at first marriage and age at first birth by at least 0.4 and 0.3 years respectively [31]. The reviews also implied that stipend programs have a significant positive impact on the girls’ productivity as well as wage earnings [31], and can increase female empowerment through positive marriage market outcomes in the long-term [25].

Due to sustained investment in the education sector and continuation of education transfer programs like FSSP, Bangladesh increased access to primary and secondary education and has succeeded in providing equal access to girls at primary and secondary level [32]. Data from routine education monitoring system show that since Bangladesh’s independence in 1971, number of schools more than tripled, and the number of students enrolled in primary and secondary levels increased by 351% and 555% respectively (see Figure 1) [33, 34].

**Figure 1.**
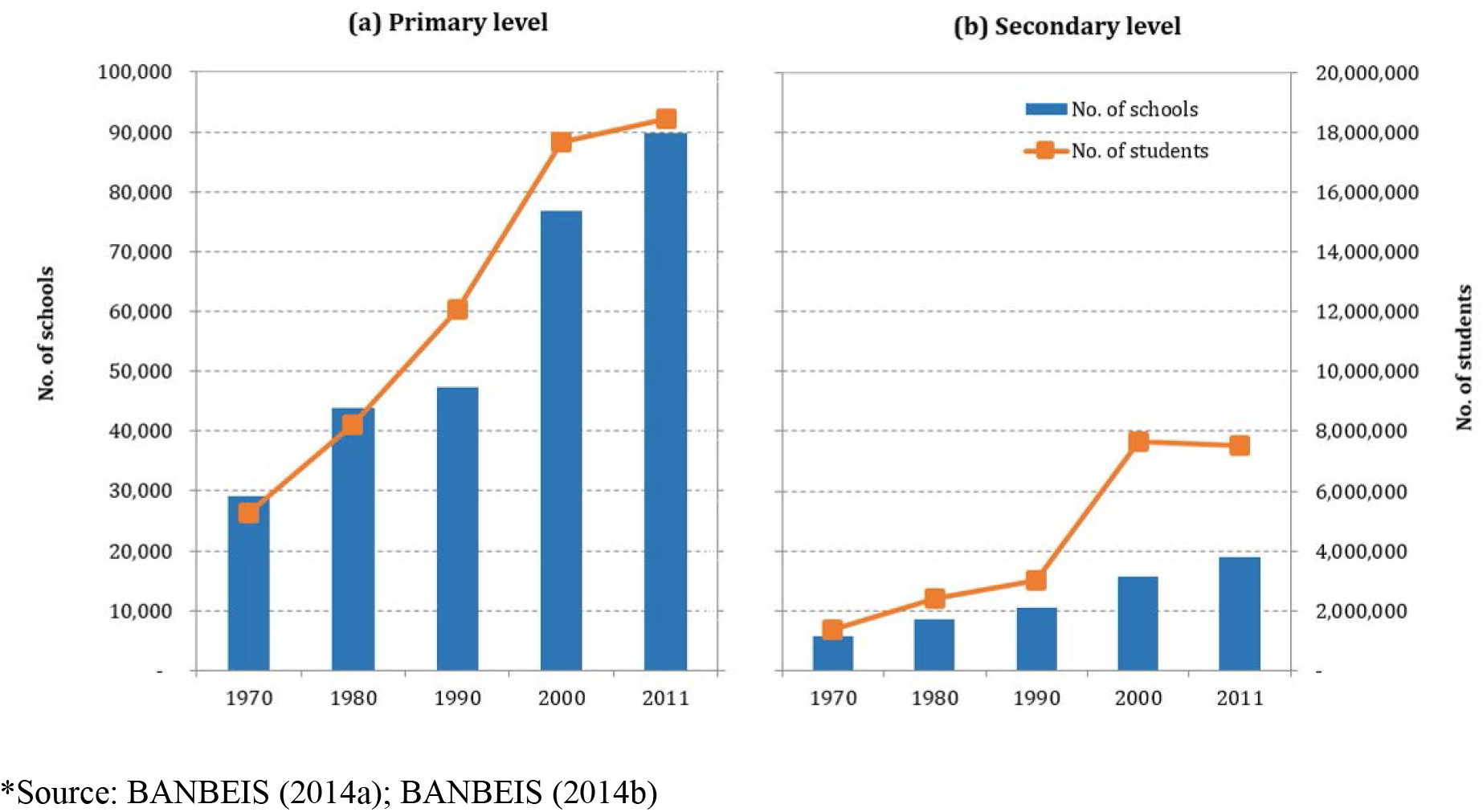
Education institutes and enrolment in Bangladesh, 1970–2011*

During 1993–2011, educational attainment among ever-married women of reproductive age (i.e. 15–49 years) has changed drastically—the proportion of women with no education declined from 58% to 28%, whereas proportion of women with secondary incomplete or higher education increased from 15% to 42%. Proportion of women with primary complete or lower education stayed around 30% during this period (see Figure 2) [23, 24].

**Figure 2.**
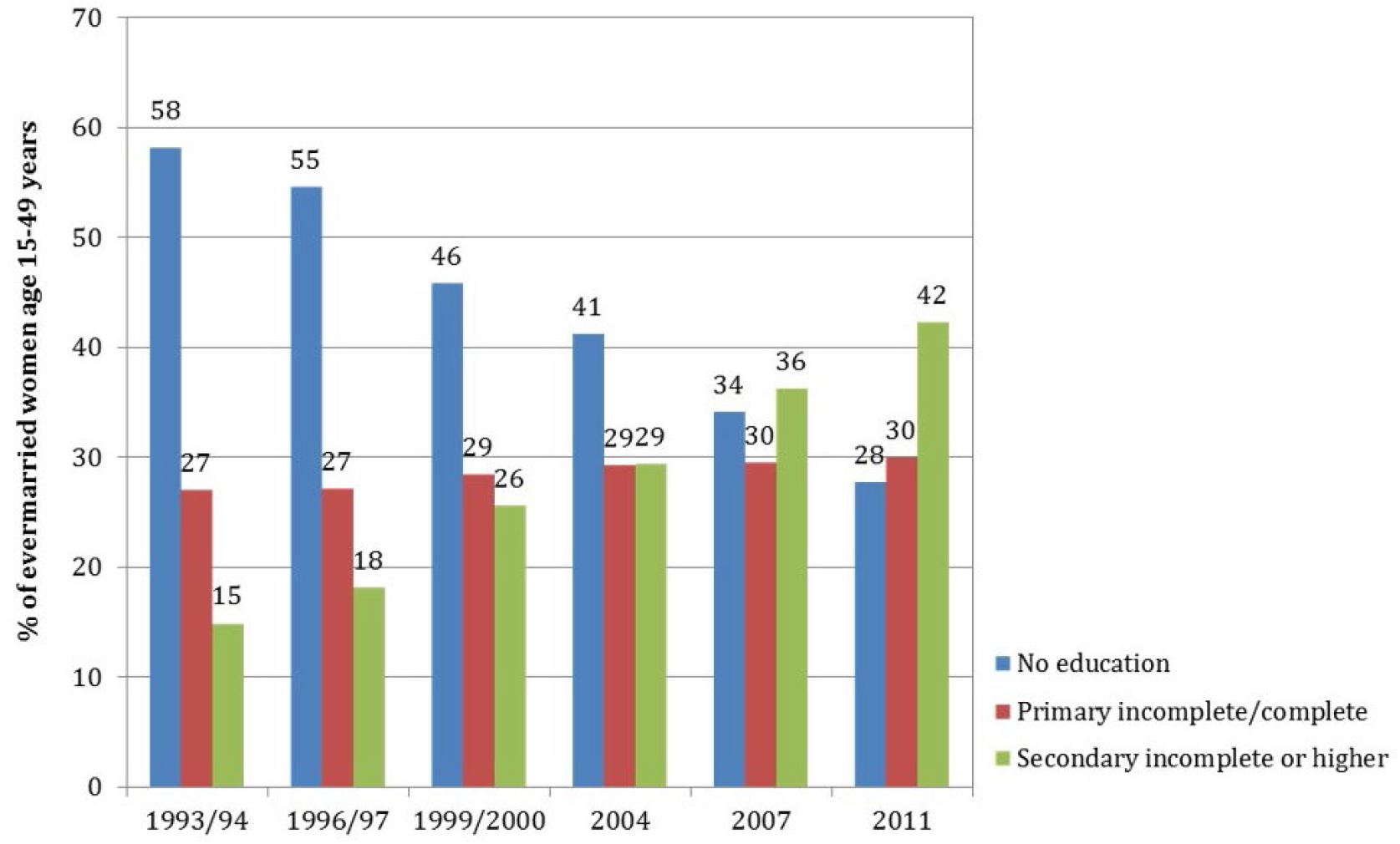
Educational attainment among ever-married women of reproductive age in Bangladesh, 1993/94–2011

## Data and Methods

In order to estimate the role of education levels in averting maternal deaths up to 2030, projected figures for women of reproductive age (WRA) are divided into broad education levels based on current trends, and MMR by educational attainment is applied to projected WRA figures. The estimated maternal deaths were decomposed to determine the role of improved education status among WRA and other factors (viz. fertility) and compared to 2030 EPMM target set for Bangladesh to determine the future course of action.

Bangladesh is unique among low- and middle-income countries in having two nationally-representative and high-quality household surveys focused on maternal mortality conducted since the year 2000. Data from these two surveys, the 2001 and 2010 Bangladesh Maternal Mortality Surveys (BMMS), were used for estimating MMR by education levels for this study. Both surveys were large, covering approximately 100,000 and 174,000 households respectively, and used the same methodology (multi-stage sample selection procedure) to generate nationally representative estimates for maternal mortality ratio. All deaths among women of reproductive age (13–49 years) in the sampled households within three years preceding the survey rounds were subsequently followed up using a verbal autopsy questionnaire to assess cause of death. To maximize comparability, both the survey rounds used a similar verbal autopsy protocol that comprised both structured and unstructured questions to collect information from the most knowledgeable household member regarding the woman’s death. The BMMS verbal autopsy tool included more detailed questions associated with pregnancy and delivery than the 2007 World Health Organization (WHO) verbal autopsy tool [4, 5].

The projected fertility levels and population figures for Bangladesh over the next two decades were taken from the ‘World Population Prospects: The 2012 Revision’ by the United Nations [35]. The World Population Prospects have been providing population estimates and projections for all countries of the world since the 1950s, and these estimates are being used by international organizations as well as academic researchers as a standard input for development planning, monitoring and global modelling [36]. The 2012 Revision of the World Population Prospects used probabilistic methods for projecting fertility and mortality, along with incorporating the results of the most recent round of national population censuses and findings from recent specialized demographic surveys that have been carried out around the world [35].

As the focus of this study is WRA, education attainment among ever-married women of reproductive age from six BDHS rounds was used. The BDHS is also based on a multi-stage sample selection procedure and generate nationally representative estimates of background characteristics (e.g., age, education, religion, media exposure) and reproductive health care practices of ever-married WRA residing in non-institutional dwelling units in the country [24].

Exponential trends were fitted to the progression in major educational attainment levels (i.e. no education, primary incomplete/complete, and secondary incomplete or higher) among WRA during 1993–2011 in order to project education attainment up to 2030. For projection of educational attainment among WRA, we considered two scenarios: a) scenario I where we assumed a conservative rate of progress for 2011–2030, slightly lower than the trends observed during 1993–2011, considering the substantial increase in WRA in the coming decades that may put pressure on the absorption capacity of primary and secondary schools in Bangladesh; and b) scenario II where we assumed that education levels would continue to improve among WRA in line with the observed rate during 1993–2011.

The equation used for estimating maternal deaths from the projected population, fertility, educational attainment, and MMR was as follows [37]:

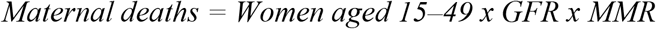

The number of maternal deaths averted in 2030 through improved education levels was estimated by applying 2010 MMR levels for different educational attainment to the proportion of ever-married women of reproductive age in Bangladesh throughout the study period (i.e. 2011–2030). The total number of maternal deaths averted between 2011 and 2030 was calculated as the difference between (a) the number that would have occurred had educational attainment remained unchanged since 2011 (effect of fertility); and (b) the estimated 2011 number when education levels changed in the context of rising female education under both the scenarios.

## Results

Analysis of data from 2001 and 2010 BMMS shows that MMR varies considerably by education level. In 1998–2001, MMR among women with no education was 370 deaths per 100,000 live births and among women with secondary or higher education was 187 deaths per 100,000 live births. Variation in MMR was more pronounced in 2007–2010, with MMR of 439 deaths per 100,000 live births among women with no education and 90 deaths per 100,000 live births among women with secondary or higher education.

Although population growth rates are slowing in Bangladesh, there is still a major component of population momentum built in to the system, so numbers of females (and males) in the reproductive ages will continue rise through the mid-21^st^ century and peak at close to 204 million around 2060 [35]. It can be seen that although the number of woman of reproductive age will increase during 2001–2030, births will fall considerably (from 3.5 million to 2.7 million annually), bringing down the General Fertility Rate (GFR) from 107 to 54 births per 1,000 women aged 15–49 during the same period (see Table 1).

**Table 1.**
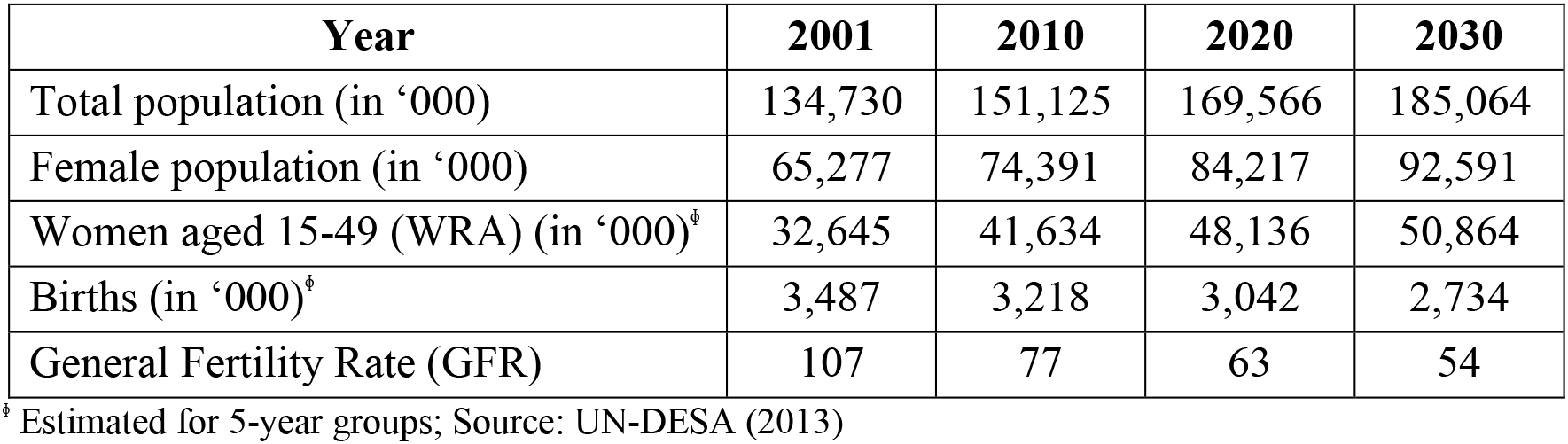
Projected demographic trends in Bangladesh, 2001–2030.

To project educational attainment among WRA under the scenario I (i.e. conservative estimate for progress in female education for 2011–2030), we used an exponential model. The exponential trend lines fitted on proportions of ever-married WRA indicate that by 2030, around 12% of women will have no education, 28% will have primary education, and 60% will have secondary incomplete or higher education (see Figure 3a). The coefficients of determination (R^2^) of the fitted curves were 0.98 (for proportion of women with no education), 0.92 (for proportion of women with primary education), and 0.96 (for proportion of women with secondary or higher education), which demonstrate a high goodness of fit. On the other hand, we used a linear model to project educational attainment among WRA under the scenario II (i.e. assuming that education levels would continue to improve in line with the observed rate during 1993–2011). The fitted trend lines under scenario II indicate that by 2030, less than 5% of women will have no education, 27% will have primary education, and 70% will have secondary incomplete or higher education (see Figure 3a). Like the previous scenario, the coefficients of determination (R^2^) of the fitted curves under scenario II for all educational attainment levels also remain 0.95 or higher, demonstrating a high goodness of fit.

**Figure 3.**
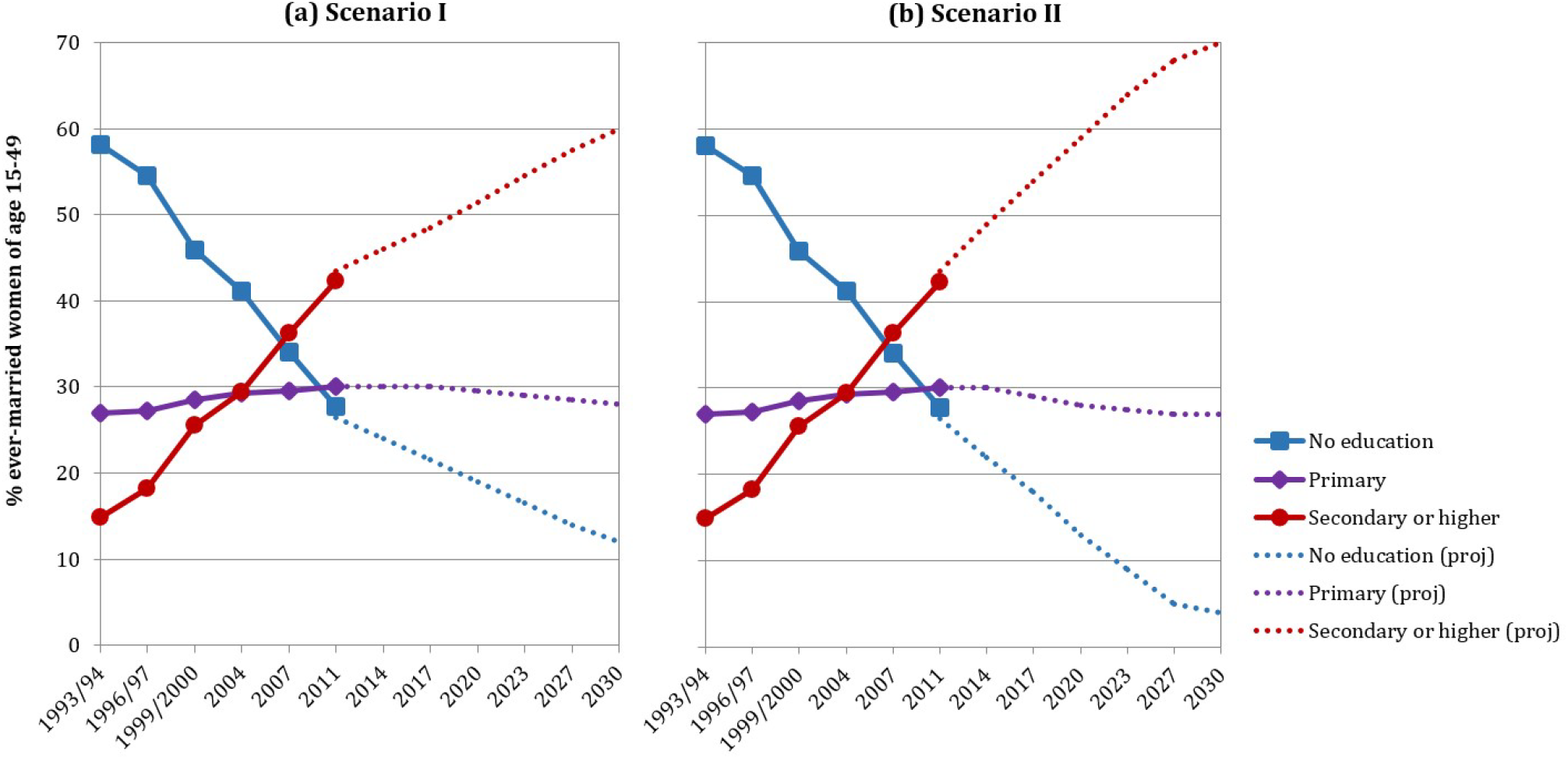
Projected trends in educational attainment among women in Bangladesh, 1993/94–2030

Based on the 2011 distribution of educational attainment among ever-married WRA in Bangladesh under the scenario I, we estimated that the total number of maternal deaths in Bangladesh would have been 5,917 in 2030 (15% lower than 6,964 maternal deaths in 2011) if educational attainment had remained unchanged. This in fact captures the role of fertility change (i.e. fewer births) in averting maternal mortality. The number of maternal deaths in the context of rising education levels among WRA in 2030 was estimated to be 4,471 (corresponding to an MMR of 164 deaths per 100,000 live births), i.e., 24% of the deaths can further be averted only by improving the female education levels over time at a slower rate than the observed trend. However, in order to achieve the EPMM target for Bangladesh of an MMR of 59 deaths by 100,000 live births by 2030 (i.e. 1,612 maternal deaths in a year), a further 64% reduction will be required (see Figure 4-a).

**Figure 4.**
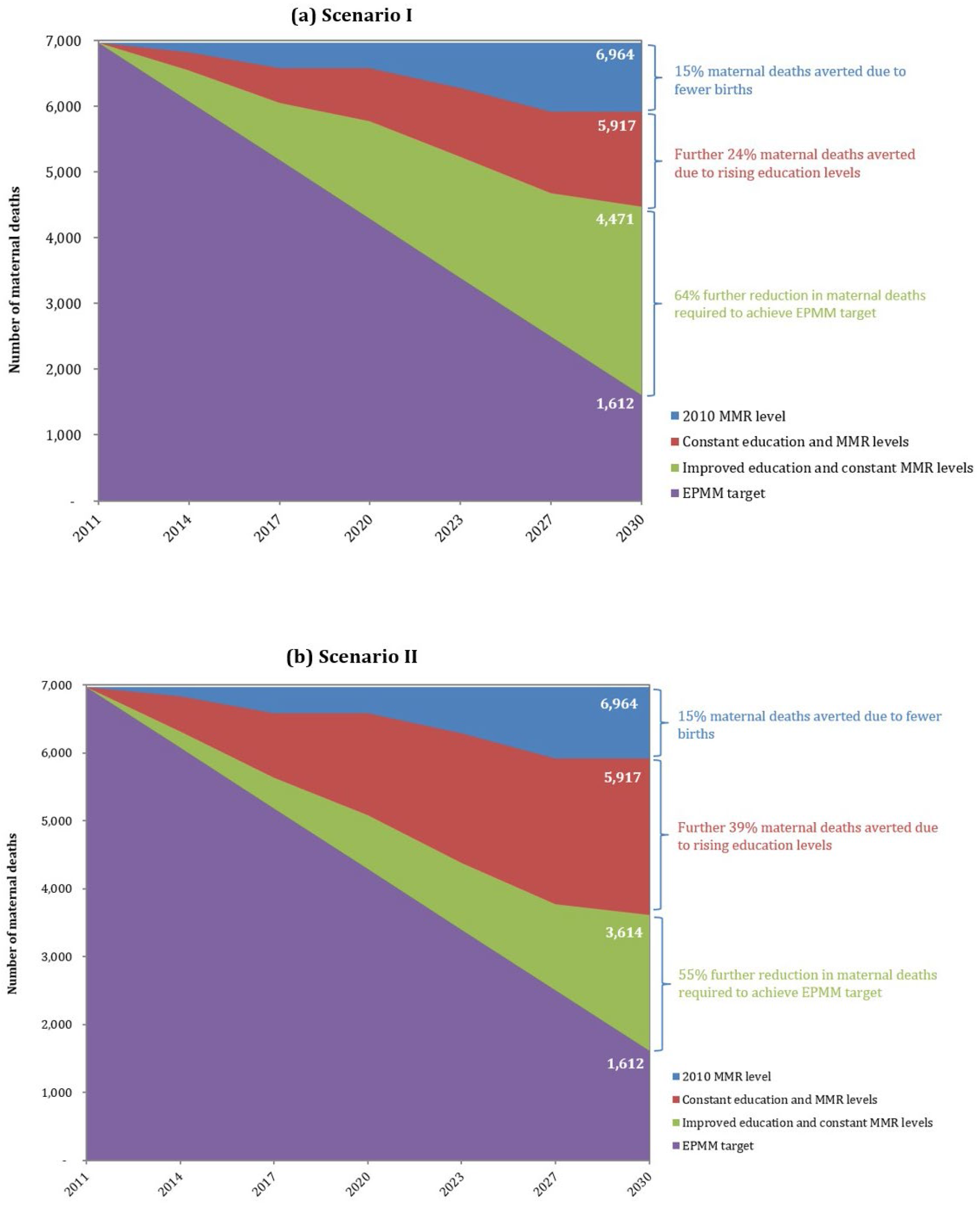
Maternal deaths in the context of rising education levels in Bangladesh, 2011–2030

Under the alternative scenario (scenario II) where the rise in educational attainment among WRA continues as per the trend observed during 1993–2011, then the reduction in maternal deaths by 2030 due to improved education levels will be 39%. In that case a further 55% reduction will be required to achieve the EPMM target in 2030 (see Figure 4-b). This scenario suggests that even with a very optimistic outlook, progress in fertility decline and female education over the next decade will bring Bangladesh only halfway towards the EPMM target by 2030.

## Discussions and Policy Considerations

Female education is considered to be a highly effective intervention to increase age at marriage and first birth, and can potentially lead to empowering women and improving nutrition [18, 25, 38]. Evidence from developing countries shows that supply side incentives (e.g. stipend) have a positive impact on human capital formation by increasing household income and access to information for informed decision making [39, 40]. This paper estimated reduction in maternal mortality based on the assumption that female educational attainment will continue to improve over the next decade. Review of female education levels over time in a number of low- and lower-middle income countries like Armenia, Indonesia, Vietnam, Ghana and Bolivia indicates that with strong political focus on social development and investments in the education sector, it is possible to bring down the proportion of women with no education to under 5% (and those with secondary or higher education to above 60%) of the female population [41]. Reaching the goals of universal access in primary and higher enrolments in secondary education in Bangladesh can be pursued through: (i) identifying hard-to-reach populations (including urban slums) to address their educational needs; (ii) scaling up stipend schemes; (iii) working in close partnership with the non-government sector; and (iv) enhancing efficiency of the education sector by better utilization of funds and improving transition rates [42].

In addition to continued investment for female education, a 64% reduction in maternal mortality to reach EPMM target can only be attained by increased and accelerated investments in improving access to and quality of health care facilities to provide care for maternal complications and safe delivery services in Bangladesh. Currently, comprehensive emergency obstetric care (CEmOC) services (needed for saving mother’s life in case of serious maternal complications) are available in 132 (out of 425) Upazila Health Complexes (UHC), 63 (out of 97) Maternal and Child Welfare Centres (MCWC), 59 District Hospitals, 3 General Hospitals and 22 public sector Medical College Hospitals [43]. However, in reality, only 25% of District Hospitals, 18% of UHCs, and 4% of MCWCs have CEmOC in terms of availability of five Obstetric First Aid items [44], and except for the medical college hospitals, none provide true 24/7 services. Given the experience with trying to keep all these facilities functional, a strong case can be made to focus the investment on a smaller number of facilities, strategically located to ensure good coverage with ease of access, and providing actual 24/7 services which will require the equipment, support and at least four full sets of staff, including obstetricians and anaesthetists. Also in terms of specific interventions, EmOC services need to ensure availability of misoprostal or oxytocin and magnesium sulphate, and rapid blood transfusion options to address the two most common obstetric causes of deaths: haemorrhage and eclampsia. The last is a critical need because less than 20% of UHCs and MCWCs have functioning transfusion facilities [44].

The Government of Bangladesh piloted a maternal voucher scheme in 21 sub-districts in 2008, which was scaled-up in 53 sub-districts in 2010, with the aim to increase utilization of maternal health care services by the poor. Under this program, a voucher entitles its holder to receive free maternal health care services including ante- and post-natal check-ups, safe delivery and treatment of complications (including caesarean sections and assisted vaginal delivery) from designated health facilities or skilled providers in the community, along with cash incentives and transport subsidies. The maternal voucher scheme in Bangladesh has shown that use of basic emergency obstetric care (BEmOC) services in UHCs can be increased several-folds and can reduce inequities in maternal health care services [45, 46, 47]. To this, we add that the primary provision of BEmOC services should be through sub-district hospitals (i.e. UHCs) and such demand-side financing schemes should focus on low-performing areas instead of national expansion, due to the high costs associated with the voucher program [47].

Though there has been a steady increase in facility delivery during recent years, almost 70% of deliveries in Bangladesh still take place at home [24]. In order to provide skilled assistance for home deliveries, the government initiated a plan in 2004 to bring on board 13,000 community skilled birth attendants (CSBA) by 2010. By 2010, less than half of this number had been trained and deployed to assist in home deliveries—however, the overall coverage of the CSBAs was only 0.4% of all deliveries in 2010 [5]. There have been recent reports of much greater success (i.e., higher coverage) with the private/NGO sector CSBAs who typically serve populations of about 3,000 in selected program areas [48]. Nevertheless, we are of the opinion that service deliveries through these options are, by nature, difficult to support and monitor, and that the low volume of services will not permit adequate skill retention (physicians and nurses, for example). We do, however, acknowledge that there will be areas in a sub-district from where it will not be convenient for women to travel to the sub-district hospitals. Home-based deliveries by CSBAs may serve these hard-to-reach areas best. A more selective use of CSBAs and UHFWCs will also allow the program to provide them with more effective technical support and back-up.

More than half of the facility deliveries took place in private sector in 2011, where cost of services is markedly higher than that in the public or NGO sectors. BMMS 2010 found that median expenditure for deliveries without complications was 14,903 Taka (US $215) in private facility, compared with 4,060 Taka (US $59) in public facilities and 2,865 Taka (US $41) in NGO facilities [24]. There are also efforts to upgrade public sector union-level (usually serving populations of 30,000) facilities, called Union Health and Family Welfare Centers (UHFWCs), to provide 24/7 normal delivery services by one trained reproductive health worker [43]. In order to progress towards the goal of universal health coverage, the government needs to focus on increasing normal deliveries in UHFWCs by CSBAs and newly trained midwifes to increase the share of public sector in maternal health delivery in the coming years.

In summary, following areas need to be prioritized for reducing maternal mortality in Bangladesh to reach EPMM target by 2030:

a. Ensuring universal access in primary and higher enrolments in secondary education, with a special focus on females and marginalized population groups;
b. Improving readiness of strategically located CEmOC facilities to provide quality 24/7 services;
c. Expanding demand-side financing schemes through UHCs to increase the primary provision of BEmOC services in low-performing areas;
d. Focusing on hard-to-reach areas to promote home-based deliveries by CSBAs, instead of national expansion;
e. Ensuring service provision and readiness of UHFWCs for normal delivery by CSBAs and midwifes to increase the share of public sector in maternal health services delivery.

## Conclusions

The country has achieved remarkable reductions in maternal mortality by investing heavily in female education and achieving reductions in fertility. This analysis attempted to estimate the impact of rising education levels on maternal mortality following the described education-maternal mortality pathways, and did not factor in the other factors affecting MMR, namely birth intervals, employment, age at marriage or first birth, etc. Education is an underlying structural determinant of maternal mortality, not a proximate determinant, having impact of most of the factors above. The estimated maternal deaths in this analysis are likely to be on the higher side, as there should be a strong cohort effect of education—as Bangladesh gets closer to universal primary education, women with no education will be older and consequently have higher risk of maternal death.

On the backdrop of the post-MDG goals and strategies, this study reiterates the necessity of investment for social development, particularly for female education, in the coming years. In order to achieve the EPMM target, Bangladesh will also need to prioritize and implement specific health sector interventions to sustain the progress in maternal health in the coming decades.

## Data Availability

The documentation and survey reports are publicly accessible, and the data files are available upon request by registered UNC/MEASURE Evaluation Dataverse users.

https://dataverse.unc.edu/dataset.xhtml

## Acknowledgement

We are grateful to the Carolina Population Center and its NIH Center grant (R24 HD050924) for general support. The study was supported by the UK Department for International Development through ICDDR,B and by the United States Agency for International Development (USAID) under the terms of MEASURE Evaluation cooperative agreement AID-OAA-L-14-00004. MEASURE Evaluation is implemented by the Carolina Population Center, University of North Carolina at Chapel Hill in partnership with ICF International; John Snow, Inc.; Management Sciences for Health; Palladium; and Tulane University. The maternal mortality estimates in the BMMSs were produced by Han Raggers of the Netherlands. We acknowledge the contribution of Nitai Chakraborty of Dhaka University, Bangladesh, in preparing the working data files for the analysis. We also thank the institutions and individuals who had made the BMMS 2001 and 2010 possible. Both the surveys were implemented under the authority of the National Institute of Population Research and Training (NIPORT) of the Government of the People’s Republic of Bangladesh with funding by the Government of the People’s Republic of Bangladesh, USAID Bangladesh, and AusAID. The data from both the surveys were collected and processed by Associates for Community and Population Research (ACPR) and Mitra and Associates. The authors’ views expressed in this publication do not necessarily reflect the views of the United States Agency for International Development (USAID) or the United States government.

